# Deep Learning-Aided Diagnosis of Autoimmune Blistering Diseases

**DOI:** 10.1101/2021.11.27.21266845

**Authors:** Daniel Cai, Abbas Roayaei Ardakany, Ferhat Ay

## Abstract

Autoimmune blistering diseases (AIBDs) are rare, chronic disorders of the skin and mucous membranes, with a broad spectrum of clinical manifestations and morphological lesions. Considering that 1) diagnosis of AIBDs is a challenging task, owing to their rarity and heterogeneous clinical features, and 2) misdiagnoses are common, and the resulting diagnostic delay is a major factor in their high mortality rate, patient prognosis stands to benefit greatly from the development of a computer-aided diagnostic (CAD) tool for AIBDs. Artificial intelligence (AI) research into rare skin diseases like AIBDs is severely underrepresented, due to a variety of factors, foremost a lack of large-scale, uniformly curated imaging data. A study by Julia S. et al. finds that, as of 2020, there exists no machine learning studies on rare skin diseases [1], despite the demonstrated success of AI in the field of dermatology. Whereas previous research has primarily looked to improve performance through extensive data collection and preprocessing, this approach remains tedious and impractical for rarer, under-documented skin diseases. This study proposes a novel approach in the development of a deep learning based diagnostic aid for AIBDs. Leveraging the visual similarities between our imaging data with pre-existing repositories, we demonstrate automated classification of AIBDs using techniques such as transfer learning and data augmentation over a convolutional neural network (CNN). A three-loop process for training is used, combining feature extraction and fine-tuning to improve performance on our classification task. Our final model retains an accuracy nearly on par with dermatologists’ diagnostic accuracy on more common skin cancers. Given the efficacy of our predictive model despite low amounts of training data, this approach holds the potential to benefit clinical diagnoses of AIBDs. Furthermore, our approach can be extrapolated to the diagnosis of other clinically similar rare diseases.

## INTRODUCTION

Autoimmune blistering diseases (AIBDs), or autoimmune bullous disorders, are rare, chronic disorders of the skin and mucous membranes with poor prognosis in the absence of treatment [2,3]. AIBDs are generally divided into four subdivisions based on level of skin affected: pemphigus, pemphigoid, IgA-mediated bullous dermatoses, and epidermolysis bullosa acquista [3]. Our research centers around the two most common manifestations: pemphigus and pemphigoid. Given their rarity and heterogeneous clinical features, pemphigus and pemphigoid lesions are often mistakenly attributed to other more common conditions, leading to considerable diagnostic delay [3,4]. For example, pemphigus vulgaris (PV), accounting for 70% of all pemphigus cases, affects only 1-5 patients per million per year and has a diagnostic delay that ranges from months to years [5,6]. Bullous pemphigoid (BP), representing for 70% of all pemphigoid cases [7], affects roughly 30 patients per million per year in the US and has a mean diagnostic delay of 6 months [8,9].

This diagnostic delay is a primary factor in their poor prognosis: PV has a 5-year mortality rate of 90% without medication, compared to 5-15% with treatment, and BP has been reported to be associated with morality rates ranging from 6 to 41% within the first year after diagnosis [10]. AIBDs diagnosis are also generally invasive, typically relying upon perilesional biopsy [4]. Considering these attributes of AIBDs, and given the rapid advances in artificial intelligence (AI) in the field of computer vision, demonstrating the utility of an AI-based CAD is of great significance in improving both patient diagnosis and treatment.

Although abundant research has demonstrated preliminary success of AI in tasks such as distinguishing skin cancers [11-13], much of this research is dependent upon the availability of large, uniform, transparent imaging datasets. Similar datasets have been compiled and made available for more common skin conditions such as melanomas. However, for rarer under-documented skin diseases like AIBDs, a lack of transparency and uniformity amongst repositories means that their data sets are lagging far behind those of well-researched conditions.

Leveraging the clinical and visual similarities between AIBDs and skin cancers, however, we can use these larger datasets to help train classification algorithms for rarer conditions such as AIBDs. Techniques such as transfer learning and data augmentation allow us to artificially inflate the pool of training data, potentially achieving a significant classification accuracy. In tandem with recent advances in computer vision, this strategy allows for a less computationally expensive model, and offers a different framework towards future development of CADs for less well-documented diseases.

## METHODS

### Data Collection and Preprocessing

Our research focuses on three major classes: pemphigus, pemphigoid, and common differential diagnoses, which comprise the first layer of the taxonomy. Within these three major classes, there exist eight total disease manifestations, comprising the second layer of the taxonomy. The differentiation between pemphigus and pemphigoid is based on the disease’s layer of manifestation in the skin, while the differentiation in the second layer is based upon the structural proteins that are attacked [14]. This tree-structure taxonomy is illustrated in Figure 1 along with sample images which demonstrate the difficulty in distinguishing between AIBDs due to their highly similar visual features.

**Figure 1.**
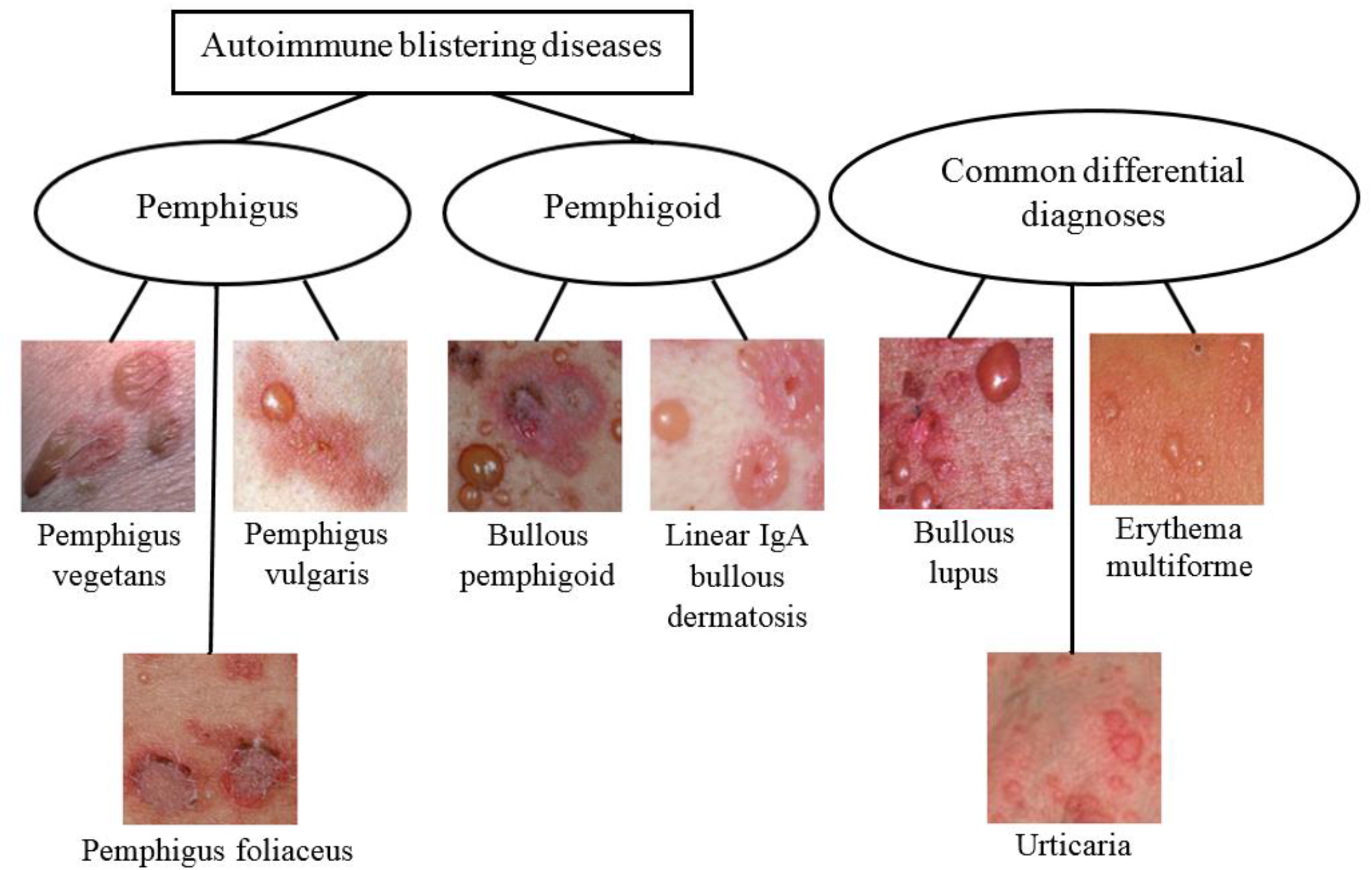
Disease taxonomy comprises of three classes in the first layer and eight classes in the second layer.

Our independently consolidated dataset consists of 1,670 clinically verified open-source images from online repositories [15-20], and accounts for various manifestations of disease ranging from oral mucosal lesions to active cutaneous blisters. The detailed dataset breakdown is listed in Table 1. Data is randomly split into training-validation-test sets, with a roughly 80-10-10 percent partitioning in order to provide the algorithm with sufficient training data.

**Table 1.**
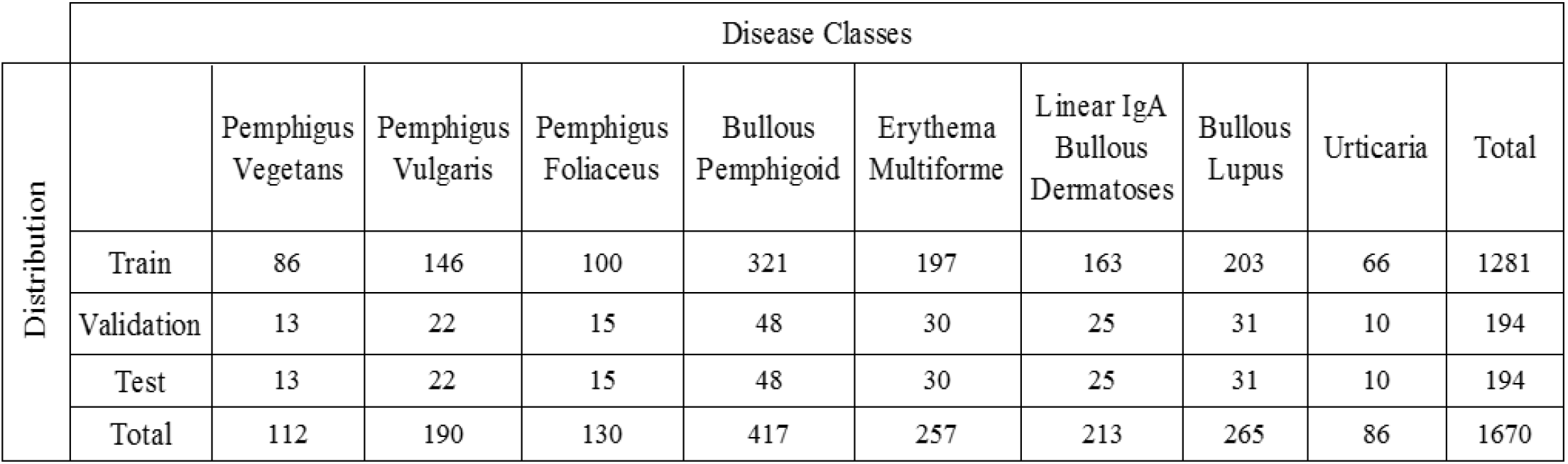
Dataset Distribution

There exist two main issues within this dataset: insufficient size and uneven class distribution. To combat these pitfalls, we prepare three distinct datasets for training:

1. 1,281 images from our main dataset
2. 2,568 images using augmented data from our main dataset Here, we augment our initial dataset with randomized rotations, scaling, and reflections of undersampled classes until we reach an equal distribution of classes. This augmentation helps combat the biases introduced by our uneven class distribution.
3. 3,000 images of skin cancers from the International Skin Imaging Collaboration (ISIC) repository The ISIC classification task is ternary, distinguishing between equal distributions of melanomas, nevi, and seborrheic keratoses.

Regularization, including image resizing and color normalization, has been applied to all three datasets.

### Deep Learning Architecture

Here we propose a prototype for classification of blisters using a convolution neural network (CNN). CNNs have gained increasing popularity in recent years over traditional machine learning approaches due to their efficacy in complex tasks like image recognition. CNNs use convolutional layers to generate invariant features, which are passed through several more filters to generate more invariant and abstract features. The process continues until a final feature is generated which is invariant to occlusions, allowing the system to accurately extract informative features from images without traditional manual image processing.

We implement the GoogleNet Inception v3 architecture, which is pre-trained on roughly 1.28 million images to 80 percent accuracy [21]. As illustrated in Figure 2, Inception v3 consists of factorized, small and asymmetric convolutions, regularized by an auxiliary classifier. More importantly, the use of novel inception modules allows for decreasing the computational expense while still maintaining efficient learning on multiple scales [21]. The modules are particularly useful in shrinking down computation time given the complexity of the blisters.

**Figure 2.**
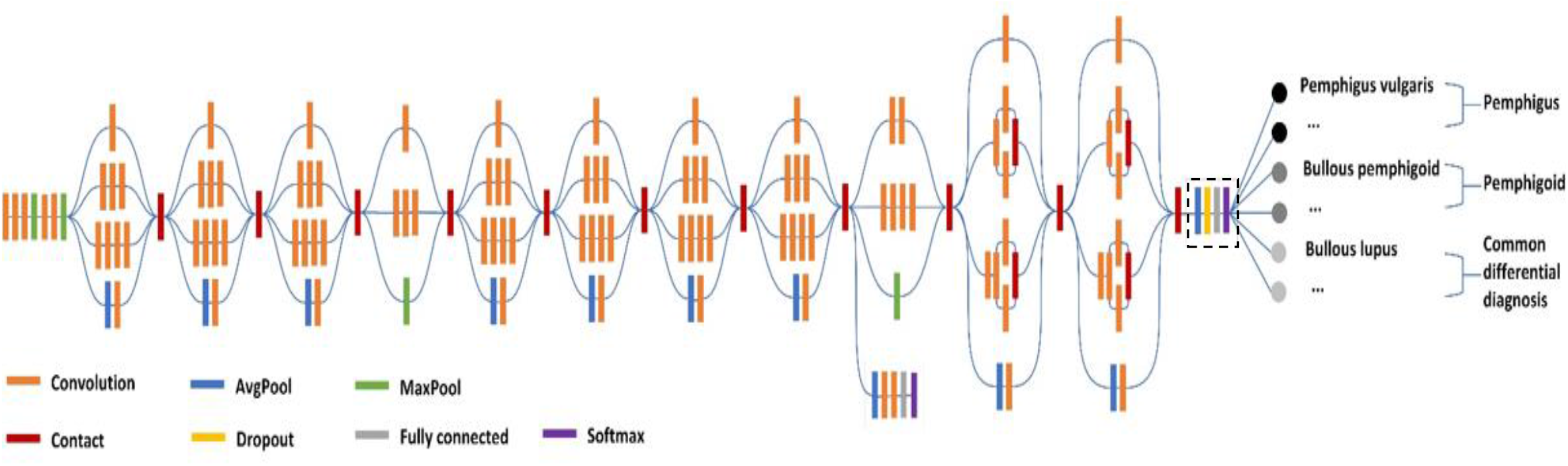
Inception v3 architecture for the task of predicting AIBDs

### Implementation of the Predictive Model

To combat insufficiencies in our training data, we employ transfer learning. In transfer learning, the model developed for one task is reused as the starting point for another task. In our case, the stored knowledge gained in both the ISIC and augmented datasets is then applied to our model for AIBD classification.

The two most common incarnations of transfer learning in the context of deep learning are:

#### Feature Extraction

By pretraining on a similar dataset, the model learns to extract certain features. As opposed to a randomly initialized CNN, therefore, our model is able to integrate this knowledge as it trains on the main dataset.

#### Fine-tuning

In contrast to training on uneven class distributions, fine-tuning trains initially on the augmented dataset. We then train on the raw, uneven dataset with a very low learning rate, allowing for the model to incrementally adapt without the risk of overfitting.

Our training consists of three progressive sequential training loops, utilizing feature extraction and fine-tuning. Between each loop, we freeze certain layers of the previously trained model, so that the new training set does not override the previous learned information. We treat the exact number of layers to freeze as a hyperparameter, initially freezing up to the fully connected layer, and then manually adjusting the threshold as illustrated in Figure 3. A full schematic of our procedure is shown in Figure 5.

**Figure 3.**
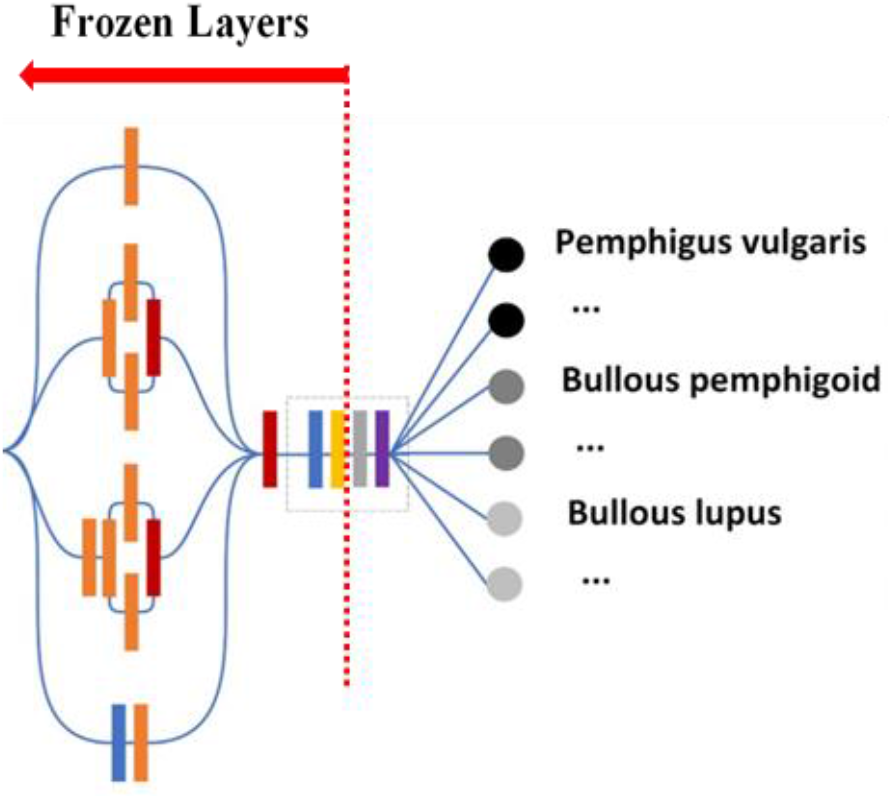
Frozen Layers/Parameters

**Figure 4.**
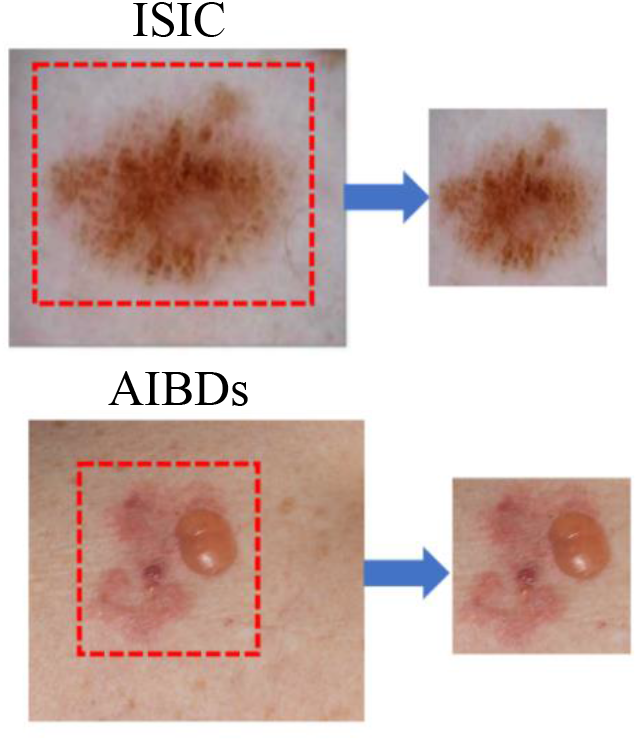
Feature extraction allows for tasks like image segmentation to be pretrained in our model

**Figure 5.**
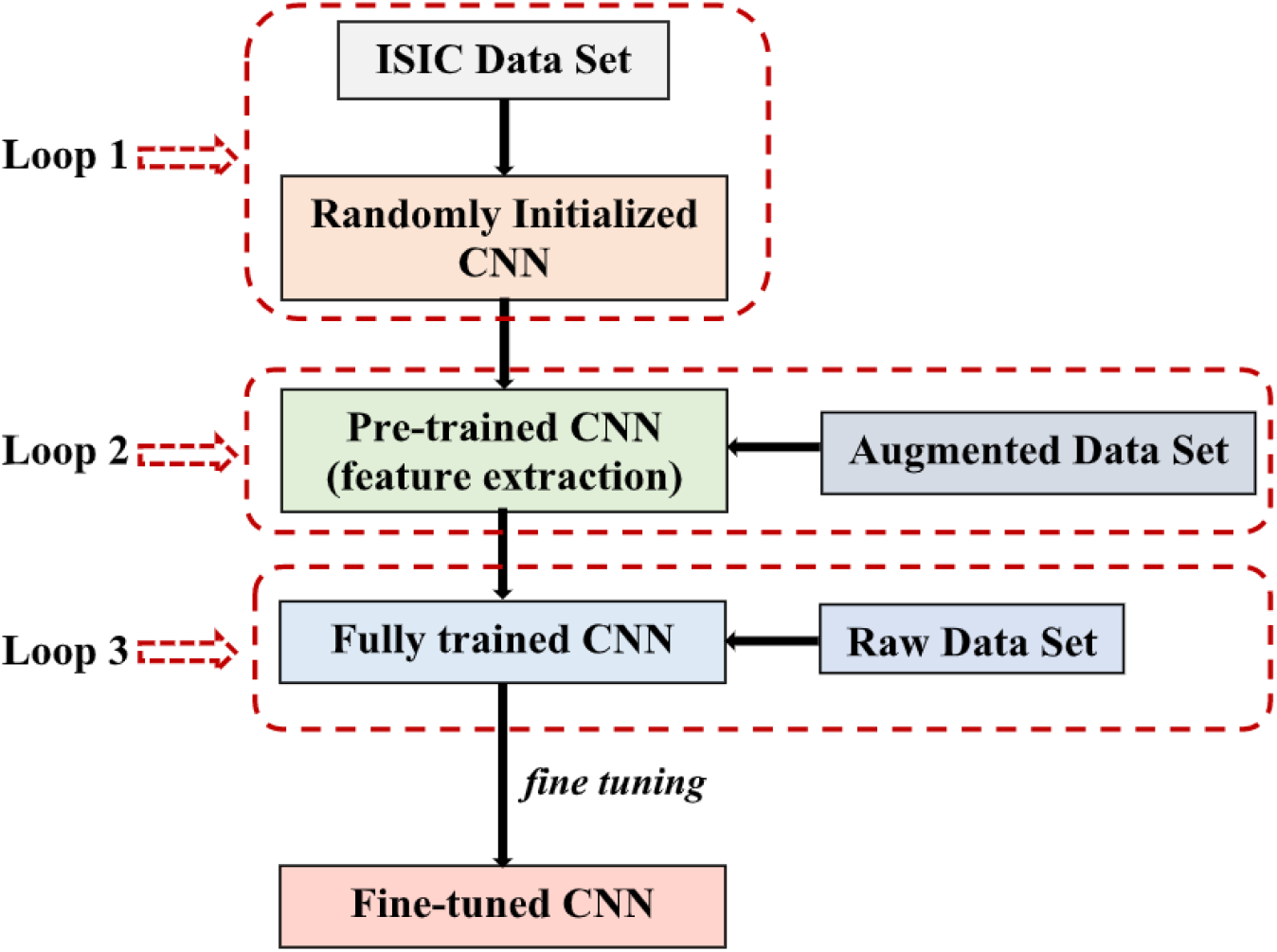
Schematic diagram of three-loop transfer learning procedure

Our first loop trains on the ISIC dataset, learning to differentiate between three types of skin cancers. The similar nature of these skin cancer conditions allows the algorithm to train on a much larger dataset and transfer its learned features over to our task. We transfer this model over as an integrated feature extractor, meaning that the model has learned to complete tasks like image segmentation (Figure 4) even before training on AIBD images.

Our second loop trains on an augmented dataset. In early trials the algorithm placed little weight on the underrepresented classes, resulting in low sensitivity (see “Metrics” section) for the majority of the disease classes. We thus augment these underrepresented classes to introduce a bias that counteracts this imbalance. Our goal here is not to improve raw accuracy, but to ensure sensitivity is improved amongst most classes.

Our third loop trains on our original main dataset (1,281 original images from AIBDs). Our goal here is to mainly fine-tune the previous model. In addition to freezing layers, we decrease the learning rate while increasing the dropout rate to make sure any adjustments the algorithm makes are minor, not overriding previous knowledge.

Certain factors such as skin color and lighting may skew our training. To combat this, we implement early stopping within our algorithm (which halts training if validation error greatly exceeds training error) in order to prevent overfitting to these invariants.

### Metrics and Analysis Methods

We aim to report results on three main metrics:

1. Accuracy between the three main classes of the first layer (pemphigus, pemphigoid, and common differential diagnoses)
2. Accuracy between the eight subdivided classes of the second layer
3. Sensitivity of the first and second layers, according to the following definition

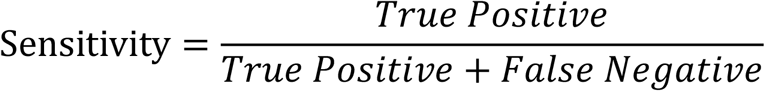

Accuracy indicates a general effectiveness of the algorithm while sensitivity allows us to address the medically significant issues of false positives and negatives.

The diagnostic accuracy of dermatologists presented in previous research is used as a reference to evaluate our diagnosis accuracy. In a 2017 study focused on AI-aided diagnosis of common skin lesions, dermatologists examined a binary benign vs malignant differentiation of skin cancers, and also a nine-class disease partition. Tasks were conducted by certified dermatologists, achieved a 66% accuracy in the binary task, and a 55% accuracy in the second, 9-class partition [22].

Unlike other common dermatological conditions, early manifestations of autoimmune disease are often not cutaneous [22], as shown in Figure 6. Paired their rarity with wide range and complexity of differential diagnoses, pemphigus and pemphigoid are more easily misdiagnosed than most skin cancers [22]. Whereas the general public is more cognizant of skin cancer, the early symptoms of autoimmune diseases are more subtle, so patients are more likely to seek help from general practitioners than certified dermatologists. Thus, our goal is to create a tool with accuracy that matches or improves upon real-world diagnostic accuracy.

**Figure 6.**
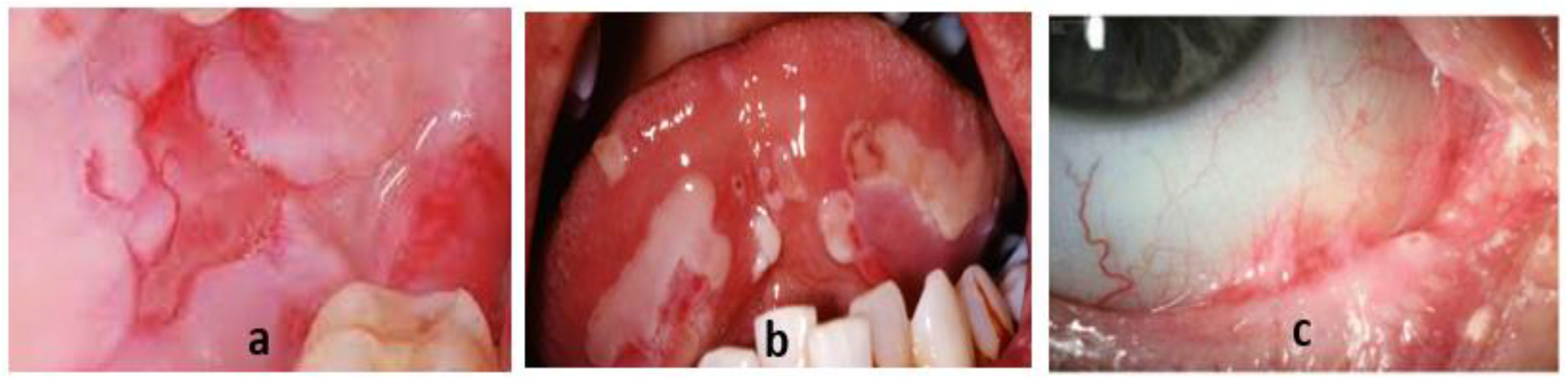
Non-cutaneous manifestations of disease. a, b: Oral lesions are common manifestations of pemphigus and pemphigus-like diseases, carrying a range of differential diagnoses including lichen planus, ulcerative stomatitis, and herpetic lesions [23-25]. c: Ocular lesions are also common manifestations of pemphigoid and pemphigoid-like diseases[26].

## RESULTS

To assess the accuracy of our trained model, a total of 194 hold out images from the eight different disease classes are tested. Our findings are summarized in Table 2 as a confusion matrix, from which the sensitivity is derived and listed in Table 3. Our CNN model achieves 67.5% accuracy on the broader disease classes (first layer), with 56.7% accuracy on the finer partitions (second layer).

**Table 2.** Raw Confusion Matrix

| True Class | Predicted Class |  |  |  |  |  |  |  |  |
| --- | --- | --- | --- | --- | --- | --- | --- | --- | --- |
|  |  | Pemphigus Vegetans | Pemphigus Vulgaris | Pemphigus Foliaceus | Bullous Pemphigoid | Linear IgA Bullous Dermatoses | Erythema Multiforme | Bullous Lupus | Urticaria |
|  | Pemphigus Vegetans | 8 | 0 | 0 | 5 | 0 | 0 | 0 | 0 |
|  | Pemphigus Vulgaris | 0 | 6 | 0 | 15 | 0 | 0 | 1 | 0 |
|  | Pemphigus Foliaceus | 0 | 8 | 4 | 2 | 0 | 0 | 0 | 1 |
|  | Bullous Pemphigoid | 2 | 2 | 0 | 40 | 2 | 0 | 2 | 0 |
|  | Linear IgA Bullous Dermatoses | 0 | 3 | 0 | 3 | 17 | 0 | 4 | 3 |
|  | Erythema Multiforme | 0 | 0 | 0 | 10 | 1 | 9 | 4 | 1 |
|  | Bullous Lupus | 2 | 0 | 0 | 5 | 0 | 0 | 22 | 2 |
|  | Urticaria | 0 | 0 | 0 | 5 | 0 | 0 | 1 | 4 |

**Table 3.** Sensitivity of both layers

| Disease Class | Sensitivity | Disease Class | Sensitivity |
| --- | --- | --- | --- |
| Pemphigus Vegetans | 0.62 | Pemphigus | 0.52 |
| Pemphigus Vulgaris | 0.27 |  |  |
| Pemphigus Foliaceus | 0.27 |  |  |
| Bullous Pemphigoid | 0.83 | Pemphigoid | 0.79 |
| Linear IgA Bullous Dermatoses | 0.57 |  |  |
| Erythema Multiforme | 0.36 | Differential Diagnoses | 0.65 |
| Bullous Lupus | 0.71 |  |  |
| Urticaria | 0.4 |  |  |

We first assess the effectiveness of transfer learning and data augmentation in our model.

Our pretrained model has a significantly lower initial validation loss, as shown in Figure 7, indicating the efficacy of feature extraction in improving the model’s performance. The slight convergence can best be explained by excess noise within the validation data set, which may have skewed the loss.

**Figure 7.**
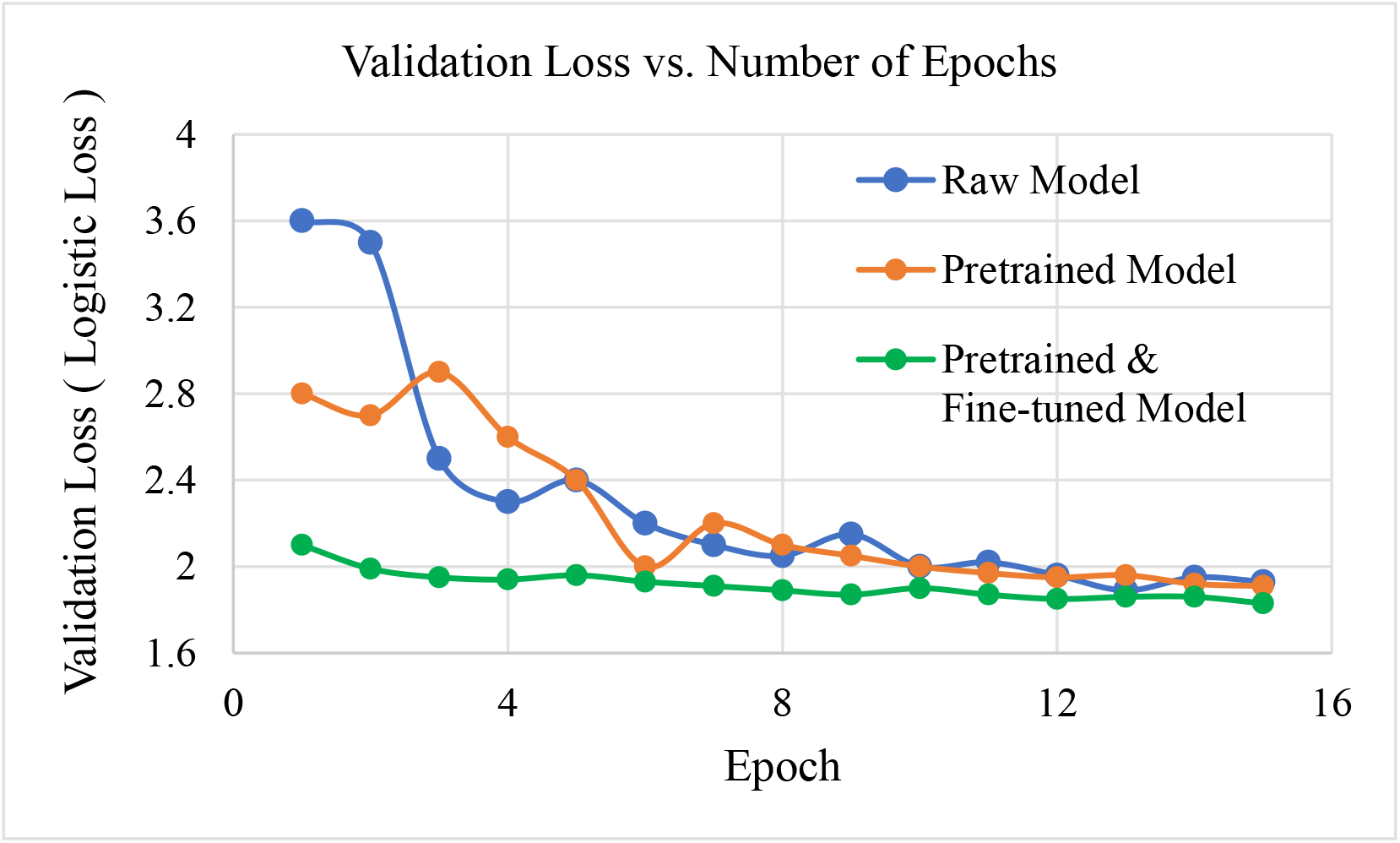
Validation loss progression of various models

However, transfer learning indeed significantly increased the training accuracy of our model. The addition of transfer learning appears to boost the training accuracy from just under 40% to almost 50% (Figure 8). Given the visual similarities between the skin cancer images in the ISIC dataset and our AIBDs image dataset, and given this increase in accuracy, it is clear that the feature extraction from the ISIC dataset effectively transfers into our AIBD imaging data.

**Figure 8.**
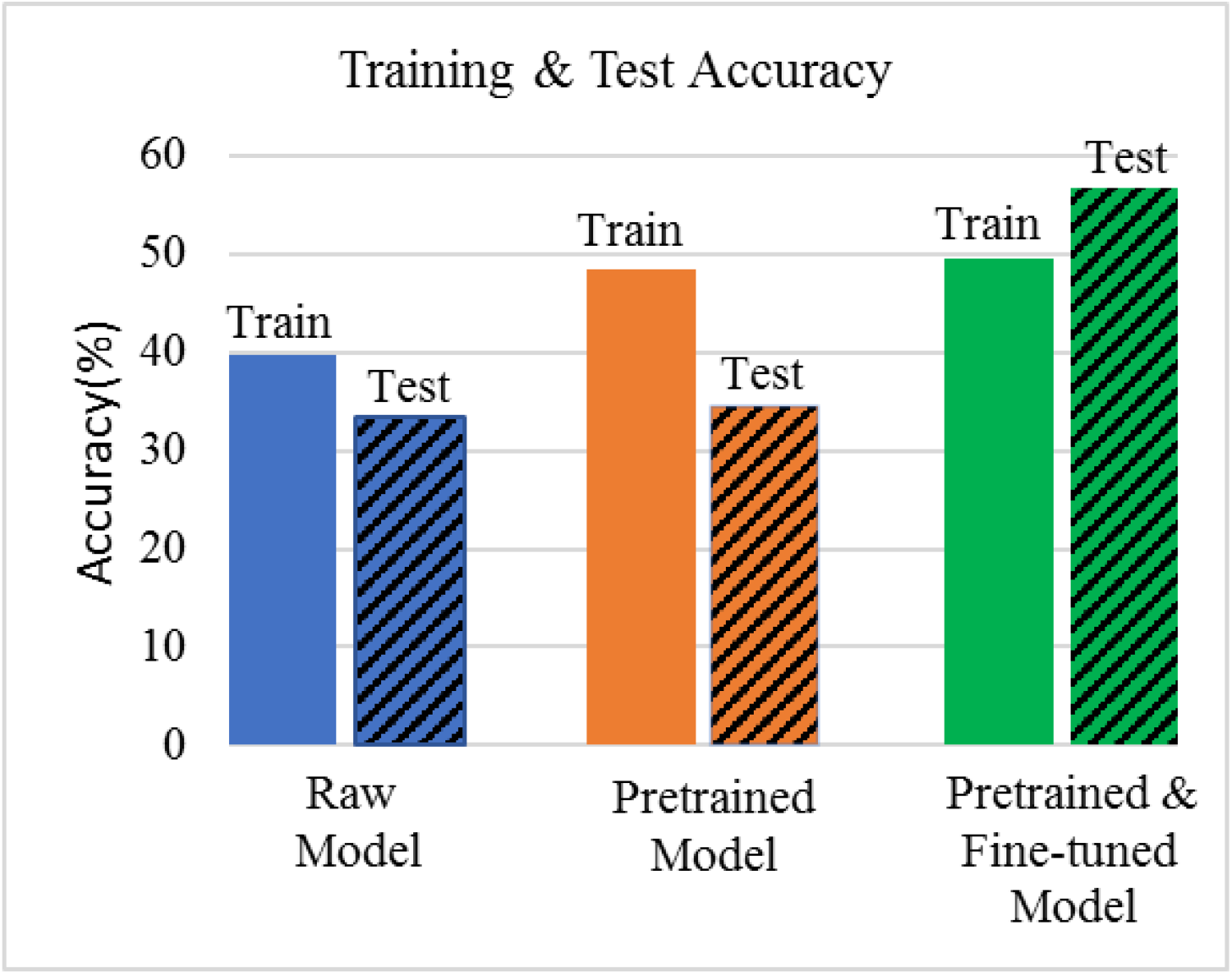
Training and test accuracy of various models

Additionally, Table 4 indicates that data augmentation and subsequent fine-tuning greatly increases the sensitivity of underrepresented classes, and Figure 8 demonstrates a significant decrease in overfitting (where training accuracy is greater than test accuracy). Our fine-tuned model (Figure 8) actually shows a reversal from the trend of overfitting, although this training-test accuracy difference is likely statistically negligible. Indeed, while the sensitivity of under sampled classes still remains below average, our model indicates a major improvement in the classification of these classes, with the sensitivity improving 2 or 3-fold over previous models (Table 4). An increase in sensitivity and decrease in overfitting reflects our model’s improved capability to generalize to other imaging data of AIBDs.

**Table 4.**
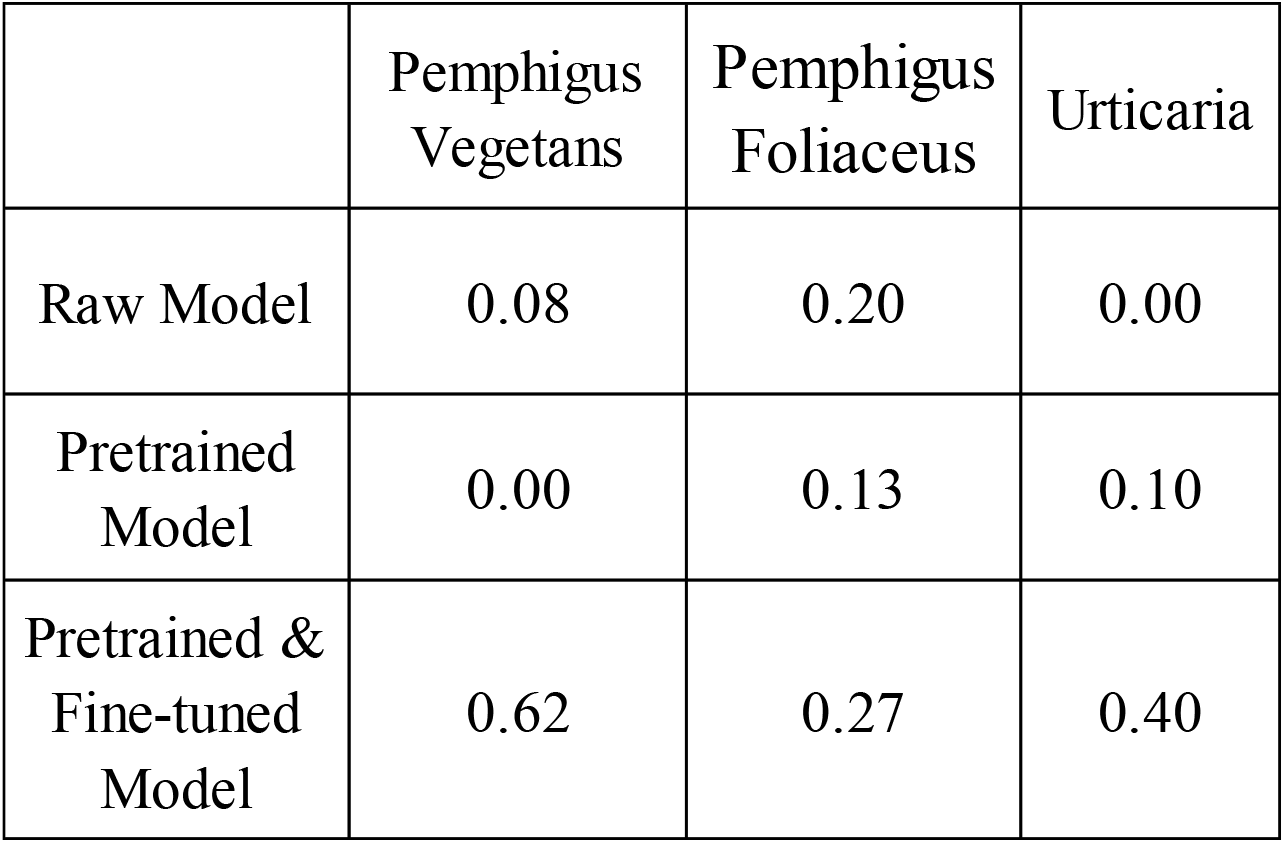
Sensitivity of undersampled classes in various models

|  | Pemphigus<br>Vegetans | Pemphigus<br>Foliaceus | Urticaria |
| --- | --- | --- | --- |
| Raw Model | 0.08 | 0.20 | 0.00 |
| Pretrained<br>Model | 0.00 | 0.13 | 0.10 |
| Pretrained &<br>Fine-tuned<br>Model | 0.62 | 0.27 | 0.40 |

Overall, the majority of classes have high sensitivity (above 65%), with the exception of the pemphigus classes which have sensitivity around 50% (Table 3). This low sensitivity is due to an uneven class distribution, which was not fully fixed through the data augmentation/oversampling. In the first layer-partition, pemphigus and pemphigoid retain high sensitivity on par with dermatologists (Table 3), indicating the effectiveness of the algorithm in differentiating between major disease classes. In a clinical setting, the high sensitivity with regards to pemphigus and pemphigoid is promising, considering the severity of false negatives for such diseases.

## CONCLUSION

Our findings demonstrate the feasibility of deep learning as an effective aid in the diagnosis of autoimmune blistering diseases. Even with a relatively small, variant dataset, through our three-loop training process incorporating transfer learning and data augmentation, our single CNN was able to classify these rare diseases with accuracy nearly on par with that of dermatologists on more common skin cancers. Our work proposes an alternative approach to deep-learning aided classification, using less computational power and requiring fewer imaging data. Given that the main constraint to this procedure is visual similarity between skin cancers and selected disease classes, this approach can easily be extrapolated to other dermatological conditions.

With a growing amount of dermatology image repositories – most notably, the ISIC and HAM10000 – and their growing disease diversity, transfer learning and data augmentation are imperative in expanding the scope of the imaging data available. While our preprocessing augmented data using conventional functions, more complex functions such as MixUp [27] may potentially better alleviate issues of overfitting and memorization. Specific to our algorithm, implementing cross-validation will improve the algorithm’s ability to generalize.

To be successfully extrapolated to other dermatological conditions, ideally our method must also be able to identify the exact features – both concrete and abstract – that were most predictive of the different AIBDs. This leads us to the concept of DeepLIFT, a novel decomposition method that allows us to make better light of the “black box” nature of neural networks [28]. With a better understanding of feature importance, it will be easier to interpret the predictive performance of the model in different scenarios, which will aid potential utilization of our tool in a clinical setting.

## Data Availability

All data produced are available online at
https://www.wikidoc.org

https://dermnetnz.org

https://atlasdermatologico.com.br

https://www.dermaamin.com

https://dermatoweb.net

https://www.dermis.net

